# Illicit Fentanyl Use Independently Predicts HCV Seroconversion Among a Cohort of People Who Inject Drugs in Tijuana and San Diego

**DOI:** 10.1101/2024.03.22.24304609

**Authors:** Joseph R Friedman, Daniela Abramovitz, Britt Skaathun, Gudelia Rangel, Alicia Harvey-Vera, Carlos F Vera, Irina Artamonova, Sheryl Muñoz, Natasha K Martin, William H Eger, Katie Bailey, Bo-Shan Go, Philippe Bourgois, Steffanie A Strathdee

## Abstract

**Background:** Illicitly manufactured fentanyl (IMF) increases overdose mortality, but its role in infectious disease transmission is unknown. We examined whether IMF use predicts HCV and HIV incidence among a cohort of people who inject drugs (PWID) in San Diego, CA and Tijuana, Mexico.

**Methods:** PWID were recruited into a prospective cohort in two waves during 2020-2022, undergoing semi-annual interviewer-administered surveys, HIV and HCV serology through February 2024. Cox regression was conducted to examine predictors of seroconversion considering self-reported IMF use as a fixed or lagged, time-dependent covariate.

**Results:** Of 398 PWID at baseline, 67% resided in San Diego, 70% were male, median age was 43, 42% reported receptive needle sharing and 25% reported using IMF. Participants contributed a median of 6 semi-annual study visits (IQR:4-6). HCV incidence was 14.26 per 100 person-years (95% CI: 11.49-17.02), and HIV incidence was 1.29 (1.00-2.28). IMF was associated with HCV seroconversion, with a univariable hazard ratio (HR) of 1.68 (95%CI: 1.12-2.53) which remained significant in multivariable models (adjHR1.54; 95%CI:1.01-2.34). The direction of the relationship with HIV was similar, albeit not significant, with an HR of 2.53 (0.7-9.15).

**Conclusion:** We document a novel association between IMF and HCV seroconversion among PWID in Tijuana-San Diego. There was insufficient power to detect if a similar relationship held for HIV. IMF’s short half-life may destabilize PWID— increasing the need for repeat dosing and sharing smoking materials and syringes. Tailoring medication dosing for opioid use disorder and new preventative care approaches may reduce HCV transmission in the fentanyl era.

**Summary:** In this cohort study of people who inject drugs in Tijuana, Mexico, and San Diego, California, fentanyl use was independently associated with HCV seroconversion. Tailored treatment and prevention efforts are needed for patients using fentanyl to minimize blood-borne infections.

## Introduction

Illicitly manufactured fentanyls (IMF) have transformed the risk environment for people who use drugs (PWUD) in North America [1]. Having outcompeted heroin for dominance in the illicit opioid market, IMF have caused dramatic increases in overdose mortality over the past decade [2]. Given their high potency, seemingly small fluctuations in product quality can result in large increases in overdose risk [1].

IMF also have additional properties that may elevate infectious disease transmission risk, including shorter half-lives and more powerful euphoric effects relative to heroin and other opioids [3]. This has been associated with increased injection frequency to prevent withdrawal symptoms and achieve sustained effects [4]. IMF use has consequently been associated with higher-risk injection practices compared to heroin use, such as sharing syringes [5–7]. Literature describing implications of IMF use, including drug preparation and administration practices [8] and behavioral aspects (including the pursuit of euphoria) [9], suggest that there may be an important—albeit not currently described—link between IMF use and human immunodeficiency virus (HIV) and hepatitis C virus (HCV). Sharply rising HCV incidence among young people over the past two decades has been linked to increasing rates of injection drug use nationally, although the particular role of fentanyl has not been determined [10–12].

We examined whether IMF use predicts HIV and HCV incidence among a longitudinal cohort of people who inject drugs (PWID) in the US-Mexico border region of Tijuana, Mexico, and San Diego, California, a region with a large population of vulnerable PWID who experience a high burden of HCV, and where IMF has broadly overtaken the drug supply.

## Methods

### Study Design and Participants

Participants were drawn from the *La Frontera* study, which is a prospective cohort of PWID in Tijuana, Mexico and San Diego, California, focused on HIV, HCV and overdose. Eligible participants were age 18+, reported injection drug use in the past month (confirmed by inspection of injection marks), spoke English or Spanish, and resided in San Diego or Tijuana.

Data were collected by trained bilingual interviewers, using a mobile outreach van targeting areas with concentrated injection drug use. Recruitment occurred in two waves from October 2020-October 2021 and February 2022-June 2022 (Supplemental Table 1). The parent study aimed to examine the role of cross-border mobility on infectious disease transmission and thus specifically recruited half of the San Diego resident sample to be individuals who reported crossing the border to use drugs in Tijuana in recruitment wave one. All wave two participants were San Diego residents. Participants underwent semi-annual interviewer-administered surveys, as well as HIV and HCV serology, through February 2024 and received $20 USD for each study visit. Study activities were approved by Institutional Review Boards at the University of California San Diego and Universidad Xochicalco in Tijuana. All participants provided written informed consent.

For this study, 720 participants (612 in wave one and 108 in wave two) were initially assessed. To assess HCV incidence, 280 individuals testing HCV-seropositive at baseline were excluded as well as two individuals with missing HCV serology. Of the 438 who tested HCV-seronegative at baseline, 398 (90.9%) completed at least one follow-up visit and comprised the analytic sample. Of 40 individuals lost to follow-up, 10 (25%) were reported as deceased, of which six were confirmed. Similarly, 35 participants who tested HIV-seropositive at baseline were excluded from HIV incidence calculations.

### HCV and HIV Serology

At each semi-annual study visit, HCV and HIV serostatus were assessed by rapid immunoassays based on blood samples [13,14] that were approved for use in the US or Mexico. Participants in San Diego were first administered a Medmira® Miriad combined HIV/HCV immunoassay (sensitivity [se]:79%-88%, specificity[sp]:100%). For any participants testing seropositive, a second line of rapid tests were conducted with Orasure® HIV (se:99.3%-100%,sp:100%) and HCV (se: 97%-98%,sp:100%). In Tijuana, Accutrak® HIV (se: 100%, sp: 100%) and HCV (se:100%, sp: 97-99%) tests were used for all participants. Participants with a reactive first rapid test underwent a second line of rapid testing with Intec® for HIV and Quality® for HCV, respectively. All samples that were reactive at the second line of rapid testing were sent to UCSD Laboratories for ELISA-based confirmatory testing.

### Survey Measures

Sociodemographic and behavioral characteristics assessed at baseline and 6-month intervals included age, sex assigned at birth, ethnicity, city of residence, housing status, substance use behaviors, and others. For each 6-month period, we assessed if participants had knowingly used fentanyl, heroin, methamphetamine, or cocaine via self-report. For the purposes of this study, fentanyl use was operationalized as both a fixed baseline characteristic and a time-varying characteristic (with a 12-month lag) to reflect the temporal association between fentanyl use and subsequent increases in infectious disease transmission risks reported in the literature[5].

### Statistical Analysis

We summarized baseline characteristics by HCV incidence status and calculated overall HIV and HCV incidence density rates per 100 person-years, as well as rates stratified by key variables chosen *a priori*, including 95% confidence intervals calculated assuming a Poisson distribution.

Limited statistical power precluded multivariable analyses where HIV seroconversion was the outcome. For HCV seroconversion, univariable and multivariable fixed and time-dependent Cox regression models were employed to assess the relationship between fentanyl use and HCV seroconversion, as well as other potential confounders and predictors based on subject-matter knowledge and previous literature—including substances used, injection behaviors known to increase HCV transmission risk, and key sociodemographic factors[5,6]. We employed a shared frailty (random effect) based on recruitment wave to control for potential intra-group correlations induced by group-specific recruitment criteria (i.e., residency and cross-border drug use) as well as differences in recruitment time between the two waves which caused time spent at risk to be dependent on recruitment wave [15]. All predictors of interest were first assessed in univariable regressions as fixed, non-time-varying predictors based on values at baseline. Next, we examined variables that yielded p-values <0.20 in the first round of models, and for which repeated measures were available, as time-varying predictors. Of those, the variable form that yielded a higher marginal log-likelihood was retained for the univariable and multivariable regression models. All variables associated with time-to-HCV seroconversion in univariable analyses at *p*≤0.10 were considered candidates for inclusion in multivariable models, controlling for age and sex (which were chosen as *a priori* control variables). Only variables that yielded a p≤0.05 in an initial multivariable model were retained in the final multivariable model. The final model was checked for multi-collinearity, interactions between covariates, proportionality of hazards, and linear relationships.

To assess the potential for retention bias, participants who were included in this analysis were compared to participants who were lost to follow-up with respect to key baseline characteristics (Supplemental Table 2). All statistical analyses were performed using SAS software version 9.4 (SAS, Cary, NC), and graphics were made using R version 4.3.1. (See supplemental methods for additional analytic details).

## Results

Participant characteristics, stratified by HCV seroconversion status, are shown in Table 1 for the 398 participants in the analytic sample (i.e., n=102 individuals who seroconverted and n=296 individuals who remained HCV seronegative during follow-up). At baseline, 67.3% resided in San Diego; 69.8% were male; median age was 42.5 years; 66.8% identified as Hispanic, Latinx or Mexican; and median years of schooling was 10.5 (i.e., some secondary school/high school) [Interquartile range [IQR]: 7.0-12.0]. Baseline characteristics revealed a highly vulnerable population, with 41.5% experiencing homelessness in the past 6 months.

**Table 1.**
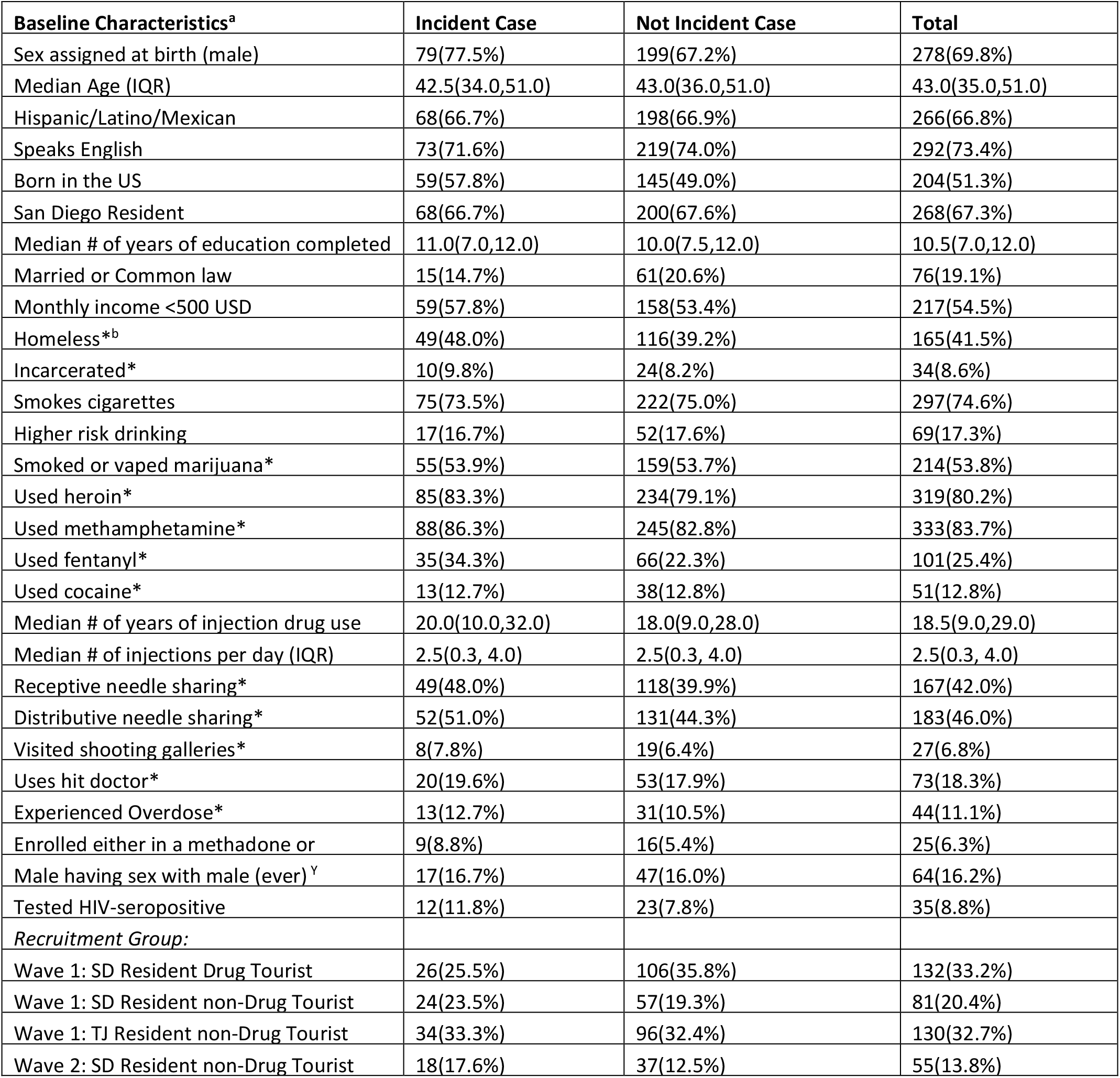
Sample characteristics at baseline for HCV incident cases vs. not incident cases. ^a^For the binary variables the affirmative category is presented; ^b^Defined based on the most common place where participants slept *Past 6 months; ^Y^Missing values n=2

Polysubstance use was common; the most commonly used substances were methamphetamine (83.7%), heroin (80.2%), cannabis (53.8%) and fentanyl (25.4%). Forty-two percent reported receptive needle sharing at baseline. The median number of average injections per day was 2.5 (IQR: 0.3-4.0). Addiction-related healthcare engagement was poor, with only 6.3% of participants reporting current enrollment in a methadone, buprenorphine or other addiction treatment program. Participants contributed a median of six semi-annual study visits (IQR:4-6).

We observed 10 HIV seroconversions during the study period (Table 2), resulting in an incidence rate of 1.29 per 100 person-years (95%CI: 1.00-2.28). Among people reporting fentanyl use, HIV incidence was 2.28 per 100 person-years (95% confidence interval[CI]: 0.05-4.50) compared to 1.00 (95%CI: 0.20-1.80) among those not reporting fentanyl use. The univariable HR for the association between fentanyl use and HIV seroconversion was 2.53 (95%CI: 0.7-9.15).

**Table 2.** HIV incidence density by past 6-month fentanyl use reported at baseline.

| Used fentanyl in past 6 months | Number of incident cases | Number of people at risk | Number of person years spent at risk | Thirty-Six Month Incidence density per 100 person years |
| --- | --- | --- | --- | --- |
| Total | 10 | 363 | 773.36 | 1.29 (0.49, 2.10) |
| No | 6 | 268 | 597.60 | 1.00 (0.20, 1.80) |
| Yes | 4 | 95 | 175.76 | 2.28 (0.05, 4.50) |

HCV incidence over the 36-month study period was 14.26 per 100 person-years (95% CI: 11.49-17.02). Among individuals who reported using fentanyl at baseline, HCV incidence was 23.7 per 100 person-years (95%CI: 15.9-31.6) compared to 11.8 per 100 person-years (95%CI: 8.97-14.63) among those not reporting fentanyl use (Figure 1). Fentanyl use, both at baseline and as a time-varying predictor (lagged by 12 months) was significantly associated with HCV seroconversion. As a time-varying predictor, fentanyl use had a univariable hazard ratio (HR) of 1.68 (95%CI: 1.12-2.53) (Figure 2) and remained independently associated with HCV seroconversion after controlling for receptive needle sharing, sex, and age (adjHR1.54; 95%CI:1.01-2.33). Fentanyl use at baseline was also significantly associated with HCV seroconversion, with a similar magnitude to the time-varying variable (see supplemental table 5). Adjusting for city of residence or cross-border mobility did not appreciably affect parameter estimates.

**Figure 1.**
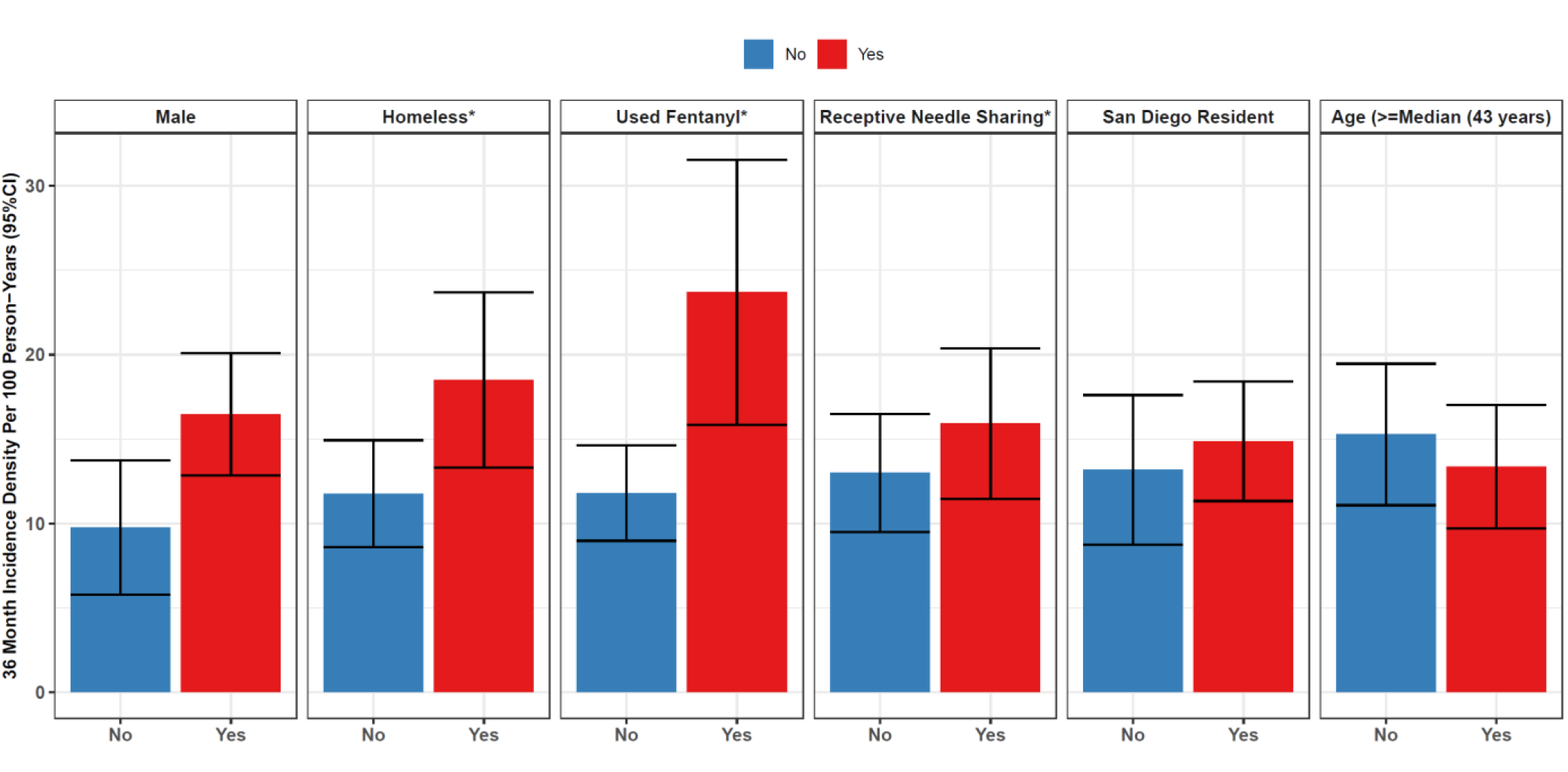
HCV Incidence Rates Stratified by Key Characteristics of Interest. HCV incidence density shown for a 36-month period per 100 person-years by key selected characteristics. Seen supplemental table 3 for exact values.

**Figure 2.**
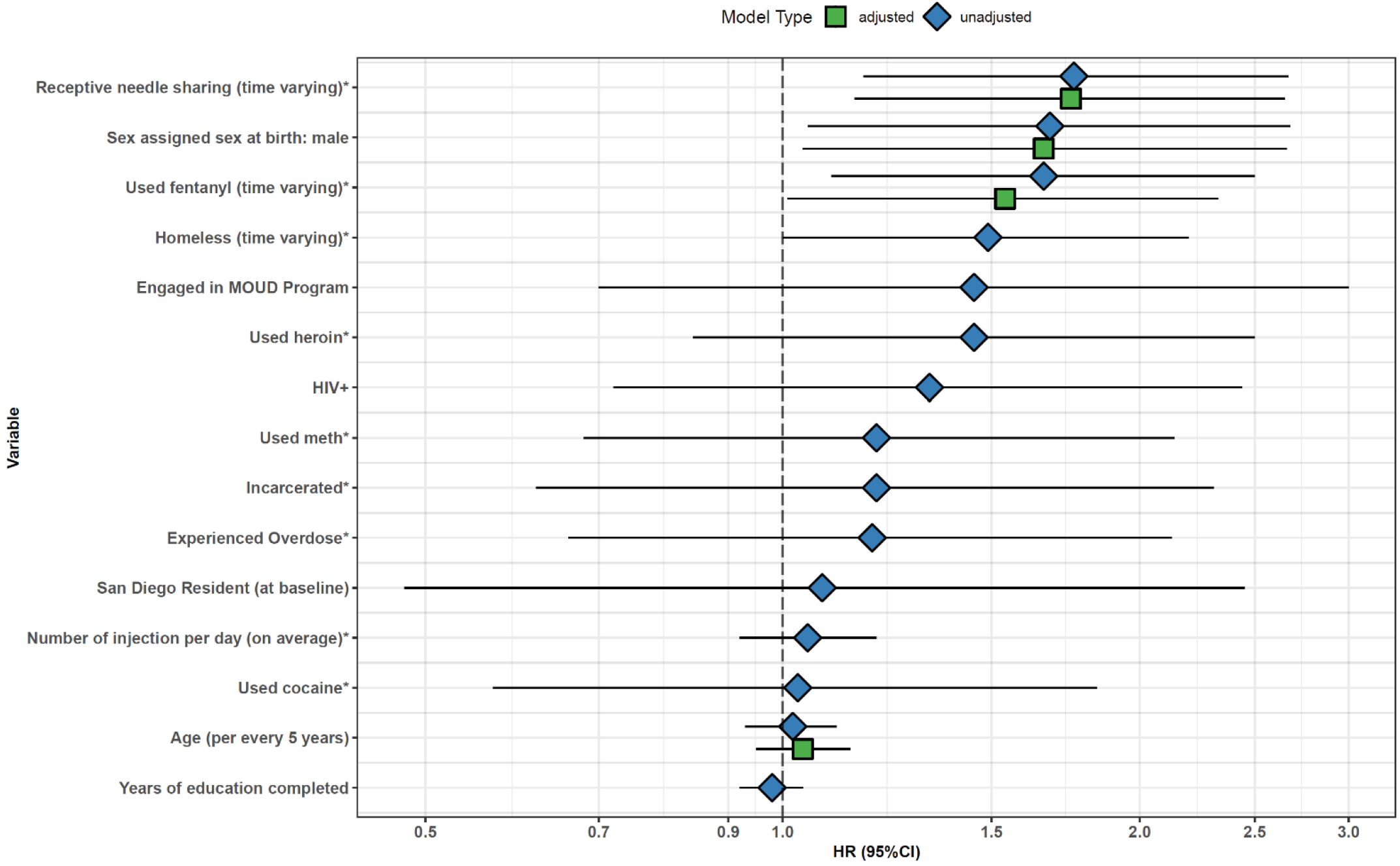
Hazard Ratios From Time Dependent Cox Models for HCV Seroconversion. Adjusted and unadjusted (univariable) model results are shown using a log scale on the x-axis. See supplemental table 4 for exact values. *predictor in the past 6 months

Individuals who practiced receptive needle sharing at baseline had an HCV incidence rate of 15.9 per 100 person-years (95%CI: 11.5-20.4) compared to 13.0 per 100 person-years among individuals who did not practice receptive needle sharing (95%CI: 9.5-16.5). In univariable models, as a time-varying predictor, receptive needle sharing was associated with a HR of 1.76 (95%CI: 1.17, 2.67), which remained significant in the multivariable model (adjHR: 1.75; 95%CI: 1.15, 2.65).

Among individuals experiencing homelessness at baseline, HCV incidence was 18.5 per 100 person-years (95%CI: 13.3-23.7) compared to 11.76 per 100-person years among those not experiencing homelessness (95%CI: 8.6-14.93). Unhoused status was associated with a univariable HR of 1.5 (95%CI: 1.0, 2.2) but its marginal association precluded us from including it in multivariable models.

Males consistently showed a higher risk of HCV seroconversion than females, with an incidence rate of 16.47 per 100 person-years (95%CI: 12.8-20.1) compared to 9.75 for females (95%CI: 5.77-13.74). Male sex had an unadjusted HR of 1.68 (95%CI: 1.05-2.68) and an adjusted hazard ratio of 1.66 (95%CI: 1.04-2.66) for HCV seroconversion. No significant differences were found for age or city of residence.

## Discussion

We document a novel association between IMF use and HCV seroconversion among PWID in Tijuana-San Diego. Although IMF have drastically transformed health risks for PWID, most research has focused on increases in fatal overdoses [1]. Less is known about the effects of the broad transition to IMF on HIV and HCV transmission. Leveraging a prospective cohort design and employing street outreach techniques to reach clandestine populations, we found a robust association between IMF use and HCV seroconversion. These findings suggest that dramatic increases in IMF use in North America since 2013 may represent a key, albeit understudied driver of contemporaneous increases in HCV incidence [11]. Given the recent White House HCV elimination plan [16], our findings suggest IMF-specific risk patterns may warrant further study to guide the implementation of the nation’s renewed efforts to reduce the burden of HCV. Although the direction and magnitude of the association between IMF use and HIV seroconversion was similar to that of HCV, the small number of HIV seroconversions precluded our ability to examine this relationship controlling for other covariates.

More research is needed to understand the mechanisms that may be underpinning the relationship between IMF use and infectious disease transmission. The shorter half-life of most IMF analogues is likely playing a key role. Qualitative and ethnographic data can help us understand the impact of this shift. Compared to the longer half-life of heroin, IMF require much more frequent dosing to prevent withdrawal symptoms, which decreases individuals’ ability to work a complete shift at their place of employment, sleep through the night, or conduct other aspects of life without continuous interruptions to use IMF [4]. This is also a frequently stated reason why PWUD seek to augment IMF with other substances, such as methamphetamine, benzodiazepines, non-IMF opioids such as nitazenes, cannabinoids, and xylazine to extend the subjective duration of effect [4,17]. These properties may account for the strong association between IMF and increased injection frequency, as well as syringe sharing that has been reported previously [5–7]. However, IMF remained significantly associated with HCV seroconversion even after accounting for receptive syringe sharing, suggesting that there may be additional effects warranting consideration in understanding the causal mechanism. It is possible that IMF’s overall destabilizing effect on the lives of PWID and associated polysubstance use may induce turbulent life circumstances, decreasing their engagement in health and preventative services and predisposing them to behaviors that increase their vulnerability to HCV infection.

An important contextual factor is the extreme vulnerability of PWID in the study region, especially in Tijuana where HCV prevalence among PWID is higher than in San Diego[18]. Social vulnerabilities represent critical drivers of HCV and HIV risk[19]. Previous studies have shown that a critical factor driving PWID to share syringes and preparation materials is a limited ability to carry paraphernalia without risking police surveillance, violence, and incarceration [20]. Municipal police have historically created challenging circumstances in Tijuana that limit PWID in their ability to carry sterile syringes [20,21]. Similar problems have also been noted in parts of California [22]. There is also a dearth of harm reduction and other services for PWID available in Tijuana, especially given recent withdrawals of governmental funding for the civil sector [21]. Harm reduction services in San Diego were also relatively limited earlier in the study period, due to the COVID-19 pandemic [28]; however, we found no difference in the weekly rates of interviews conducted inside vs. outside COVID-19 restrictive periods.

The relationship between IMF and HCV raises important implications for treatment and prevention efforts among PWID to prevent further infectious disease transmission. For instance, tailoring dosing of medications for opioid use disorder (MOUD), to respond to the unique pressures of IMF, will be important for reducing HCV transmission risks, as MOUD is known to reduce transmission risk[23]. The lipophilic nature of IMF has complicated buprenorphine induction, making precipitated withdrawal much more likely. However, novel approaches including microinduction (small buprenorphine doses scaled up as IMF are tapered), macroinduction (large starting doses of buprenorphine), and bridging to buprenorphine with short-acting opioids, all offer promising avenues to stabilize patients withdrawing from IMF [24–26]. Treatment modalities for people using IMF will require flexibility to promote MOUD uptake and adherence. For instance, mobile treatment units may connect PWID to healthcare and facilitate MOUD initiation and HCV and HIV testing[27]. Telehealth approaches may also be helpful for PWID who own electronic devices but find travelling for care difficult[28]. Long-acting MOUD also represent a powerful option to increase retention and decrease the negative health effects of return to IMF use [29]. In the inpatient setting, adequate pain control (using short-acting opioids) is increasingly required to manage pain and withdrawal symptoms related to IMF use and prevent against-medical-advice (AMA) discharges, which are currently on the rise [24,30]. Other kinds of infections affecting PWUD—such as skin and soft tissue infections—have also increased in recent years, which requires further study for treatment optimization in the fentanyl era [31,32]. Finally, the rise in IMF adds further evidence of the need to increase point-of-care testing and HCV treatment among PWUD. Even despite ongoing substance use, re-infection rates among PWID have been shown to be uncommon [33], and HCV treatment is cost-effective [34].

The IMF-HCV relationship also reinforces the need for increasing preventative and primary care services among PWUD. Research comparing the transmission of HCV and HIV suggests that HCV is the first infection to be acquired once young people start injecting, and there is generally a brief window of opportunity to prevent subsequent HIV infection [35]. Reaching individuals in this window can be accomplished via increasing access to low-barrier services that PWID need to remain safe (such as sterile syringes) alongside HIV and HCV testing [36]. On the West Coast of the U.S., there are also early signs of a broad transition from injecting to smoking IMF, which raises important considerations for blood-borne disease prevention efforts [37,38]. This shift may be helpful for reducing HCV transmission if smoking supplies are less likely than syringes to transfer HCV, although evidence on this topic is mixed [39]. Regardless, investing in safe smoking supplies is warranted to reduce the need for sharing equipment among the growing segment of PWUD smoking IMF.

This study has several limitations. Although the HCV and HIV assays employed here were highly sensitive and specific [13,14], the reliance on self-reported fentanyl use represents a limitation. Some PWUD may not know they were using IMF, given limited availability of drug checking services during the study period[40]. This could have caused misclassification, with a tendency to attenuate associations between IMF and HIV/HCV seroconversion towards the null. Follow-up studies using biological markers of fentanyl use should be conducted for confirmation. Additionally, given the known shift from injecting to smoking fentanyl occurring during the follow-up period [38], statistical power to detect an association between injection of IMF and HCV seroconversion may have been more limited. Additionally, we did not have a sufficient sample size of HIV seroconverters to assess the link between IMF and HIV, although this remains an important area for investigation. The results of this study apply to a highly vulnerable population of PWID on the US-Mexico border, and further study is needed to assess the generalizability to the wider North American context of IMF use.

## Conclusions

The broad shift among PWUD to using synthetic opioids such as IMF represents a profound change for the health risks of a broad swath of vulnerable individuals in North America. Although most research has focused on the implications for fatal overdose rates, we provide novel evidence that IMF use is associated with increased HCV incidence. This raises important considerations for treatment and prevention efforts, including access to point-of-care HCV testing and treatment, MOUD modalities and dosing, provision of harm reduction supplies and interventions to prevent subsequent HIV acquisition. More research is needed to understand potential mechanisms underpinning this association, as well as the role of IMF use on other infectious diseases, such as HIV and soft-tissue infections.

## Data Availability

The underlying data used in this study are protected as part of a cohort study of vulnerable individuals. Aggregated statistics may be requested from the authors on a case-by-case basis.

## Acknowledgements

JF and DA drafted the first draft of the manuscript. DA conducted the statistical analysis. JF created the graphical representations of data. All authors contributed to conceptualizing the study, revising the manuscript, and agreed upon the final results. AV and CV led fieldwork and data collection. SS obtained funding and supervised the study. The underlying data used in this study are protected as part of a cohort study of vulnerable individuals. Aggregated statistics may be requested from the authors on a case-by-case basis. This work was supported by the San Diego Center for AIDS Research (National Institute of Allergy and Infectious Diseases, grant P30AI036214) and the National Institute on Drug Abuse (grants R01DA049644-S1, R01DA049644-02S2, K01DA043412, 3K01DA043412-04S1, DP2DA049295, and T32DA023356). JRF was supported by the National Institute of General Medical Sciences (grant GM008042). WHE received funding from NIDA T32 DA023356. BS received funding from K01DA049665 and R03 DA57142. Authors declare no conflicts of interest.

## Notes

### Competing Interest Statement

The authors have declared no competing interest.

### Author Declarations

Institutional Review Boards at the University of California San Diego, in the United States and Universidad Xochicalco in Tijuana, Mexico gave ethical approval for this work. All participants provided written informed consent.

