## Supplement for "Illicit Fentanyl Use Independently Predicts HCV Seroconversion Among a Cohort of People Who Inject Drugs in Tijuana and San Diego"

### Supplement for "Illicit Fentanyl Use Independently Predicts HCV Seroconversion Among a Cohort of People Who Use Drugs on the US-Mexico Border"

### Supplemental Methods

#### Extended Description of Statistical Analysis

Study participants who tested HCV sero-negative at baseline were included in this analysis. Those who acquired HCV during the course of the study were described with respect to baseline characteristics by using frequencies and percentages for binary variables and means and standard deviations for continuous variables. Overall incidence density rates of HCV per 100 person-years, as well as rates stratified by key variables, were calculated by taking the ratio between the number of cases and the number of person-years at risk accumulated over the follow-up period and multiplying it by 100. The corresponding 95% confidence intervals (CI) were calculated using a Poisson distribution. For the incident cases, the sero-conversion was assumed to take place mid-point between the last visit when the participant tested negative and first visit when they tested positive. Depending on whether the participant sero-converted or not during the study period, their time at risk was calculated as either the length of time between their baseline assessment and sero-conversion or the length of time between their baseline assessment and their last study visit (i.e., right censored).

Univariate and multivariable Cox regression models for fixed and time-dependent covariates with a shared frailty (random effect) based on recruitment group were used to examine the associations between each explanatory variable and time to incident HCV. The recruitment variable consisted of four groups: Wave 1 San Diego-DT, Wave1 San Diego-NDT, Wave 1 Tijuana-NDT and Wave 2 (all SD-NDT). We decided to use this variable as a frailty variable to control for potential intra-group correlations induced by group-specific recruitment criteria (i.e., residency, "cross-border drug use") as well as for the differences in the recruitment time between the two waves which caused the time spent at risk to be dependent on recruitment wave (average time at risk in wave 1=27 months and in wave 2=15 months). Furthermore, the lognormal distribution was selected to express the frailty based on optimal values of Log Likelihood and AIC obtained from fitting a series of parametric models (exponential, Weibull, gamma and lognormal) to "failure" times where frailty group was used as an independent variable. Once we selected the frailty distribution, we fitted univariate Cox Regression models with shared frailty to a series of fixed independent variables, based on the variables' baseline values. These variables were selected based on subject-matter and previous literature findings. Next, all the fixed variables that yielded p-values <0.20 in the univariate regressions and for which repeated measures were also available, were examined as time-changing covariates. Of those, the ones that yielded a higher marginal log likelihood were retained in Table 2.

The multivariable model was constructed using extended Cox regression to accommodate the inclusion of both, fixed and time-varying covariates. The strategy for building the model was based on a "purposeful selection of variables" approach (Hosmer, Lemeshow, & May, 2011) guided by subject matter, relationships among potential explanatory variables and statistical significance. All variables associated with time to incident HCV in univariate analyses at $p$ =< 0.10, were considered as candidates for inclusion in the multivariable model, while controlling for age and gender. Only variables that yielded a 5% significance level in the multivariable model were retained. The final model was checked for multi-collinearity which was ruled out upon examining values of the variance inflation factors and largest condition indexes. Additionally, interactions between covariates were assessed and ruled out.

Since we used the shared frailty Cox regression models, the proportionality of hazards was assumed to be conditional on frailty. As such, we assessed this assumption separate for each frailty group. For categorical fixed independent variables we examined plots of log-minus-log of the survival function against the time at risk variable and for continuous fixed variables we examined plots of Schoenfeld residuals against the time variable. Martingale residuals plots were used to verify the linear relationship between each continuous explanatory variable and the log hazard of acquiring HCV.

To assess the potential for retention bias, participants who were included in this analysis were compared to participants who were lost to follow-up with respect to key baseline characteristics by using Chi-Squared and Fisher's exact tests for binary variables and Mann-Whitney tests for continuous variables. An alpha level of 0.05 was used to determine statistical significance.
All statistical analyses were performed using SAS software version 9.4 (SAS, Cary, NC).

### Supplemental Results

#### Supplemental Table 1. Number of observations by visit and recruitment wave

| Semi-annual Visits (Oct 2020 and Feb 2024) | Total Number of observations | Number of observations (Wave 1)* | Number of observations (Wave 2)** |
|---|---|---|---|
| Visit 1 (baseline) | 398 | 343 | 55 |
| Visit 2 (6 months) | 373 | 322 | 51 |
| Visit 3 (12 months) | 363 | 316 | 47 |
| Visit 4 (18 months) | 352 | 311 | 41 |
| Visit 5 (24 months) | 303 | 301 | 2 |
| Visit 6 (30 months) | 283 | 283 | 0 |
| Visit 7 (36 months) | 81 | 81 | 0 |

*Wave 1 (visit 1: Oct 20-Oct 21; visit 2: May 21-May 22; visit 3: Nov 21-Nov 22; visit 4: May 22-July 23; visit 5: Nov 22-Jan 24; visit 6: Jun 23-Feb 24; visit 7: Dec 23-Feb 24)
**Wave 2 (visit 1: Feb 22-Jun 22; visit 2: Aug 22-Mar 23; visit 3: Mar 23-Aug 23; visit 4: Jul 23-Jan 24; visit 5: Jan 24-Feb 24)

#### Supplemental Table 2. LF Baseline Characteristics of PWID by lost to follow-up status (among those who tested HCV negative at baseline)

| Variable[a] | Lost to follow-up N=40 | Not lost to follow-up N=398 | Total N=438 | p |
|---|---|---|---|---|
| Sex assigned at birth (male) | 30(75.0%) | 278(69.8%) | 308(70.3%) | 0.50 |
| Median Age (IQR) | 41.0(29.0,46.5) | 43.0(35.0,51.0) | 43.0(35.0,51.0) | 0.06 |
| Hispanic/Latino/Mexican | 19(47.5%) | 266(66.8%) | 285(65.1%) | 0.01 |
| Speaks English | 37(92.5%) | 292(73.4%) | 329(75.1%) | 0.01 |
| Born in the US | 33(82.5%) | 204(51.3%) | 237(54.1%) | <.001 |

| Variable[a] | Lost to follow-up N=40 | Not lost to follow-up N=398 | Total N=438 | p |
|---|---|---|---|---|
| San Diego Resident | 36(90.0%) | 268(67.3%) | 304(69.4%) | 0.00 |
| Median # of years of education completed (IQR) | 12.0(10.5,12.5) | 10.5(7.0,12.0) | 11.0(8.0,12.0) | 0.00 |
| Married or Common law | 2(5.0%) | 76(19.1%) | 78(17.8%) | 0.03 |
| Monthly income <500 USD | 16(40.0%) | 217(54.5%) | 233(53.2%) | 0.08 |
| Median # of people in the same household (IQR)* | 2.0(1.0, 4.5) | 2.0(1.0, 5.0) | 2.0(1.0, 5.0) | 0.09 |
| Homeless* | 23(57.5%) | 165(41.5%) | 188(42.9%) | 0.05 |
| Incarcerated* | 7(17.5%) | 34(8.6%) | 41(9.4%) | 0.07 |
| Smokes cigarettes | 28(70.0%) | 297(74.6%) | 325(74.2%) | 0.57 |
| Higher risk drinking | 10(25.0%) | 69(17.3%) | 79(18.0%) | 0.23 |
| Smoked or vaped marijuana* | 24(60.0%) | 214(53.8%) | 238(54.3%) | 0.45 |
| Used heroin* | 27(67.5%) | 319(80.2%) | 346(79.0%) | 0.06 |
| Used methamphetamine* | 38(95.0%) | 333(83.7%) | 371(84.7%) | 0.06 |
| Used fentanyl* | 14(35.0%) | 101(25.4%) | 115(26.3%) | 0.19 |
| Used cocaine* | 7(17.5%) | 51(12.8%) | 58(13.2%) | 0.40 |
| Median # of years of injection drug use (IQR) | 14.5(5.0,23.0) | 18.5(9.0,29.0) | 18.0(9.0,28.0) | 0.06 |
| Median # of injections per day (IQR) | 2.5(0.3, 4.0) | 2.5(0.3, 4.0) | 2.5(0.3, 4.0) | 0.49 |
| Receptive needle sharing* | 11(27.5%) | 167(42.0%) | 178(40.6%) | 0.08 |
| Distributive needle sharing* | 10(25.0%) | 183(46.0%) | 193(44.1%) | 0.01 |
| Visited shooting galleries* | 2(5.0%) | 27(6.8%) | 29(6.6%) | 0.67 |
| Uses hit doctor* | 10(25.0%) | 73(18.3%) | 83(18.9%) | 0.31 |
| Experienced Overdose* | 4(10.0%) | 44(11.1%) | 48(11.0%) | 0.84 |
| Enrolled either in a methadone or buprenorphine program or in a rehab/drug treatment center* | 6(15.0%) | 25(6.3%) | 31(7.1%) | 0.05 |
| Male having sex with male (ever)[γ] | 3(7.5%) | 64(16.2%) | 67(15.4%) | 0.15 |
| Male having sex with male*[γ] | 1(2.5%) | 21(5.3%) | 22(5.0%) | 0.44 |
| Tested HIV-seropositive | 1(2.5%) | 35(8.8%) | 36(8.2%) | 0.17 |

[a]For the binary variables the affirmative category is presented; *Past 6 months; [γ]Missing values n=2

#### Supplemental Table 3. LF HCV Incidence Density

| Variable | Stratum | Number of people at risk | Number of incident cases | Number of person years spent at risk | Thirty Eight Months Incidence density per 100 person years (95% CIs)* |
|---|---|---|---|---|---|
| San Diego Resident | No | 130 | 34 | 257.95 | 13.18( 8.75, 17.61) |
| | Yes | 268 | 68 | 457.49 | 14.86( 11.33, 18.40) |
| Age (>=Median (43 years) | No | 194 | 51 | 333.86 | 15.28( 11.08, 19.47) |
| | Yes | 204 | 51 | 381.58 | 13.37( 9.70, 17.03) |

| Variable | Stratum | Number of people at risk | Number of incident cases | Number of person years spent at risk | Thirty Eight Months Incidence density per 100 person years (95% CIs)* |
|---|---|---|---|---|---|
| Male | No | 120 | 23 | 235.83 | 9.75( 5.77, 13.74) |
| | Yes | 278 | 79 | 479.61 | 16.47( 12.84, 20.10) |
| Homeless* | No | 233 | 53 | 450.64 | 11.76( 8.60, 14.93) |
| | Yes | 165 | 49 | 264.79 | 18.51( 13.32, 23.69) |
| Used Fentanyl* | No | 297 | 67 | 567.79 | 11.80( 8.97, 14.63) |
| | Yes | 101 | 35 | 147.64 | 23.71( 15.85, 31.56) |
| Receptive Needle Sharing* | No | 231 | 53 | 407.64 | 13.00( 9.50, 16.50) |
| | Yes | 167 | 49 | 307.79 | 15.92( 11.46, 20.38) |

Exact numerical details are provided for the incidence rates shown in Figure 1. *CIs constructed based on Poisson Distribution

#### Supplemental Table 4. Univariate and multivariable Cox models with frailty by recruitment group examining factors associated with HCV seroconversion.

| Characteristics | Hazard Ratio | 95% CI | P-value | Adjusted Hazard Ratio | 95% CI | P-value |
|---|---|---|---|---|---|---|
| Sex assigned sex at birth: male | 1.68 | ( 1.05,2.68 ) | 0.03 | 1.66 | (1.04, 2.66) | 0.008 |
| Age (per every 5 years) | 1.02 | ( 0.93,1.11 ) | 0.73 | 1.04 | (0.95, 1.14) | 0.38 |
| Hispanic/Latino/Mexican | 1.13 | ( 0.70,1.81 ) | 0.62 | | | |
| Born in the US | 1.61 | ( 0.96,2.67 ) | 0.07 | | | |
| San Diego Resident | 1.08 | ( 0.48,2.45 ) | 0.85 | | | |
| Years of education completed | 0.98 | ( 0.92,1.04 ) | 0.48 | | | |
| Married or Common law | 0.72 | ( 0.42,1.25 ) | 0.25 | | | |
| Monthly income <500 USD | 1.16 | ( 0.74,1.83 ) | 0.52 | | | |
| Homeless*[v] | 1.49 | (1.00, 2.20) | 0.05 | | | |
| Incarcerated* | 1.20 | ( 0.62,2.31 ) | 0.58 | | | |
| Higher risk drinking | 0.90 | ( 0.53,1.51 ) | 0.68 | | | |

| Characteristics | Hazard Ratio | 95% CI | P-value | Adjusted Hazard Ratio | 95% CI | P-value |
|---|---|---|---|---|---|---|
| Smoked or vaped marijuana* | 1.02 | ( 0.69,1.50 ) | 0.93 | | | |
| Used heroin* | 1.45 | ( 0.84,2.50 ) | 0.18 | | | |
| Used meth* | 1.20 | ( 0.68,2.14 ) | 0.53 | | | |
| Used fentanyl* | 1.73 | ( 1.14,2.61 ) | 0.01 | | | |
| Used fentanyl*[v] | 1.66 | (1.10, 2.50) | 0.02 | 1.54 | (1.01, 2.33) | 0.04 |
| Used cocaine* | 1.03 | ( 0.57,1.84 ) | 0.93 | | | |
| # of years of injection drug use | 1.01 | ( 1.00,1.03 ) | 0.13 | | | |
| Number of injection per day (on average)* | 1.05 | ( 0.92,1.20 ) | 0.48 | | | |
| Receptive needle sharing*[v] | 1.76 | (1.17, 2.67) | 0.007 | 1.75 | (1.15, 2.65) | 0.008 |
| Visited shooting galleries* | 1.19 | ( 0.57,2.50 ) | 0.64 | | | |
| Used hit doctor* | 1.04 | ( 0.64,1.71 ) | 0.87 | | | |
| Experienced Overdose* | 1.19 | ( 0.66,2.13 ) | 0.56 | | | |
| Enrolled either in a methadone or buprenorphine program or in a rehab/drug treatment center* | 1.69 | ( 0.85,3.37 ) | 0.13 | | | |
| Male having sex with male (ever) [Y] | 1.04 | ( 0.61,1.78 ) | 0.88 | | | |
| HIV+ | 1.33 | ( 0.72,2.44 ) | 0.36 | | | |

Exact numerical details are provided for the incidence rates shown in Figure 2. *Past 6 months; [v]Time Varying; [Y]Missing values n=2

#### Supplemental Table 5. Multivariable Cox Model With Fentanyl Use as a Fixed Baseline Covariate.

| Label | Hazard Ratio | 95% Lower Confidence Limit for Hazard Ratio | 95% Upper Confidence Limit for Hazard Ratio | Pr > ChiSq |
|---|---|---|---|---|
| Used fentanyl* | 1.61 | 1.05 | 2.46 | 0.03 |
| Receptive needle sharing*[v] | 1.70 | 1.17 | 2.70 | 0.007 |

| Label | Hazard Ratio | 95% Lower Confidence Limit for Hazard Ratio | 95% Upper Confidence Limit for Hazard Ratio | Pr > ChiSq |
|---|---|---|---|---|
| Sex at birth (male vs. female) | 1.69 | 1.05 | 2.70 | 0.03 |
| Age (per every 5 years) | 1.05 | 0.95 | 1.15 | 0.34 |

*Past 6 months; [v]Time Varying;
